# Robust Determinants of Neurocognitive Development in Children: Evidence from the Pune Maternal Nutrition Study (PMNS)

**DOI:** 10.1101/2022.02.07.22270566

**Authors:** Chittaranjan S. Yajnik, Chih Ming Tan, Vidya Bhate, Souvik Bandyopadhyay, Ashwini Sankar, Rishikesh V. Behere

## Abstract

Neurocognitive development is a dynamic process over the life course and is influenced by intrauterine factors as well as later life environment. Using data from the Pune Maternal Nutrition Study (PMNS) from 1994 to 2008, we investigate the association of in-utero, birth, and childhood conditions with offspring neurocognitive development in 686 participants of the cohort, at age 12 years. The life course exposure variables in the analysis include maternal pre-pregnancy size and nutrition during pregnancy, offspring birth measurements, nutrition and physical growth at age 12 years along with parental education and socio-economic status. We used the novel Bayesian Model Averaging (BMA) approach; which has been shown to have better predictive performance over traditional tests of associations. Our study employs 8 standard neurocognitive tests that measure intelligence, working memory, visuo-conceptual and verbal learning, and decision-making/attention at 12 years of age. We control for nutritional-metabolic information based on blood measurements from the pregnant mothers and the children at 12 years of age. Our findings highlight the critical role of parental education and socioeconomic background in determining child neurocognitive performance. Maternal characteristics (pre-pregnancy BMI, fasting insulin during pregnancy) and child height at 12 years were also robust predictors on the BMA. A range of early factors – such as maternal folate and ferritin concentrations during pregnancy, and child’s head circumference at birth – remained important determinants of some dimensions of child’s neurocognitive development, but their associations were not robust once we account for model uncertainty. Our results suggest that intrauterine influences on long term neurocognitive outcomes may be potentially reversible by post birth remediation. In addition to the current nutritional interventions, public health policy should also consider social interventions in children born into families with low socio-economic status to improve human capital.

## Introduction

Fetal and childhood growth and development are important determinants of adult health and human capital ^1–3^. Neurodevelopment is a dynamic process and is influenced by genetic and environmental factors over the life course of an individual. In this model, intrauterine (including maternal nutrition and metabolism) and post-natal environment (events at birth, childhood illness, nutrition, socio-economic environment, education) have crucial influence. This is the central theme of the concept of Developmental Origins of Health and Disease (DOHaD).

Exposure of the fetus to adverse developmental influences such as famine during pregnancy (proxy for maternal-fetal undernourishment), maternal diabetes and poor socio-economic conditions have been studied in many populations, ^4–8^ and shown to result in negative impacts on a range of health outcomes including neurocognitive outcomes ^9^. Maternal macronutrient (glucose, lipids, amino acids) and micronutrient nutrition (Vitamins B12, folate, D and C, pyridoxine, and Iron) influence offspring neurocognitive development ^10^. Low maternal folate and vitamin B12 are associated with neural tube defects ^11^, developmental delays and autistic spectrum disorders ^12^. Babies born with Low Birth Weight (LBW), Small for Gestational Age (SGA) or Intrauterine Growth Restriction (IUGR) and those born pre-term have lower neurocognitive functioning scores in childhood ^13^. In prospective follow up studies in our birth cohorts we observed lower maternal B12 status during pregnancy to be associated with impaired neurocognitive performance of the offspring at 2 years^14^ and 9 years of age^15^. Poverty can lead to poor maternal nutrition, increased maternal stress and growth restriction and stunting in the offspring ^16^. These factors have been associated with neurocognitive impairment, poorer school performance, and underachievement in adulthood which may lead to a perpetual intergenerational cycle of deprivation and impaired development^17,18^.

The Pune Maternal Nutrition Study (PMNS) investigated nutritional and physiological determinants of fetal growth in a rural environment. The children born in this birth cohort have been followed up for their physical and neurocognitive development. In this paper, we investigate the associations of *in utero* exposures (e.g., maternal nutrition and metabolism during pregnancy), birth size and post-natal factors (e.g., childhood nutrition, growth and development), as well as parental education and family socioeconomic status with offspring neurocognitive development at age 12 years. Our work is novel in two ways. First, availability of prospective phenotypic data in the PMNS allows us to investigate the association of not only initial shocks (intrauterine factors), but also subsequent remedial changes in nutritional and socio-economic status across the life course of the child (until age 12 when the child takes the neurocognitive tests). Second, our paper also introduces methodological advancements in terms of data analysis to this area of research. Instead of using a pre-selected set of “benchmark” regression models (implicitly introducing unspecified prior knowledge about the outcome process), we employ Bayesian model averaging (BMA) methods ^19^ that, instead of relying on any single regression model, produce robust estimates by assigning evidentiary weights based on the data to each model in the model space, and then taking an average of model-specific estimates using these weights. There is considerable evidence that BMA performs better than using any single model; BMA point estimators and prediction intervals have been shown to minimize mean squared error,^20^ and BMA predictive distributions have optimal performance measured according to the logarithmic scoring rule.^21^

## Methods

### Data: The Pune Maternal Nutrition Study (PMNS) birth cohort

The Pune Maternal Nutrition Study (PMNS) is a preconception birth cohort ^22^ established in 1993 at Diabetes Unit, KEM Hospital Research Centre, Pune, India. The PMNS investigated the role of maternal and paternal influences on fetal growth and has followed up the offspring to study the risk of non-communicable disease in later life. The detailed design is reported in Yajnik et al (2008)^23^. In short, 2466 women in reproductive age group were recruited from the farming communities of 6 villages near Pune. Approximately 800 pregnancies were investigated in detail (18 and 28 weeks of gestation) for maternal nutritional intake on a food frequency questionnaire,^24^ physical activity,^25^ glucose tolerance, and circulating nutrient levels (vitamins B12, folate, C and D, pyridoxine and ferritin) and 762 live births occurred from June 1994 to April 1996. The babies were measured for size at birth. Using standard definitions^26,27^ based on birth weight and gestational age, Low Birth Weight (LBW) was defined as infants born with birth weight less than 2.5 kgs, Very Low Birth Weight (VLBW) as infants born with birth weight less than 2 kgs. Small for Gestational Age (SGA) was defined as less than 10^th^ centile of weight for gestational age. Preterm was defined as infants born at less than 37 weeks gestation.

The offspring were followed up every 6 months for physical growth. Every 6 years detailed cardio-metabolic measurements were made on parents and children. At 12 years of age, 686 (90% of live) offspring were followed up for physical growth and development, cardio-metabolic risk factors, nutrition, circulating micronutrient concentrations and neurocognitive performance. Stunting in the children was defined as height less than 2 standard deviation of height for age.^28^ See Figure 1 for timelines in the PMNS and participant recruitment.

**Figure 1:**
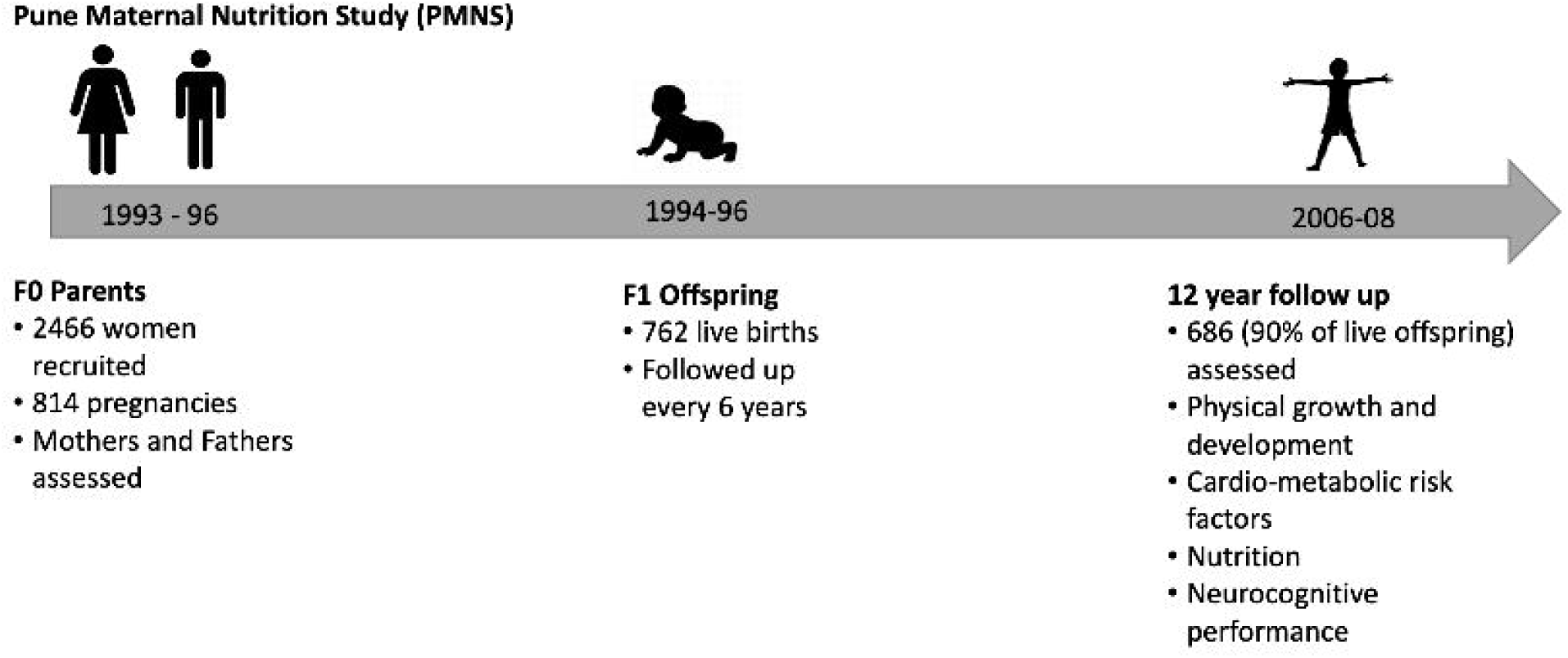
Time lines in the PMNS cohort and participant recruitment

#### Biochemical measurements

Biochemical measurements included hematological parameters, concentrations of circulating micronutrients and metabolic measurements in mothers during pregnancy and in children.^29^ Hemogram was measured on a Beckman Coulter analyzer (AC.T diffTM Analyzer, Florida, USA). Plasma vitamin B12 and red cell folate were measured using a microbiological assay technique. Total homocysteine was measured by HPLC (PerkinElmer 200 Series, PerkinElmer, Shelton, CT, USA). Plasma glucose was measured using an Hitachi 911 automated analyser (Hitachi, Tokyo, Japan) by the glucose oxidase peroxidase method. Plasma insulin was measured using a Delfia technique (Victor 2; Wallac, Turku, Finland).

#### Socioeconomic status assessment

The family’s socioeconomic status (SES) was assessed using the Pareek and Trivedi Socio economic status scale^30^. This is a validated scale for assessment of SES in the Indian rural population. It comprehensively measures SES on dimensions of caste, education, occupation, land holding, social participation, family type, type of housing, farm and material possession. A total score is derived and higher score indicates a higher SES.

#### Neurocognitive measurements

Data on our outcomes of interest were collected through neurocognitive assessments on 686 offspring at 12 years of age using 8 neuropsychological tests. The neurocognitive tests included Raven’s Colored Progressive Matrices (CPM) that measures global neurocognitive ability, Block Design (BD) test that measures visuo-spatial processing, Picture Completion (PC) test that measures visual attention, Digit Span (Forward (DF) and Backward (DB)) tests that measure attention and working memory, Color Trail Making (2 test types; TM A and TM B) tests that measure focused and divided attention, and Auditory Verbal Learning Test (AVLT) that measures verbal learning and memory (see Appendix A for a detailed description of all the tests). Data for these 8 tests were collected and Indian normative percentiles and cut-off scores were used for scoring.^31–34^ For all tests a higher score indicates better neurocognitive functioning except TM A and TM B, where higher time taken indicates a poorer outcome.

#### Ethical considerations

Written informed consent was obtained from the parents, offspring provided assent. Permission to perform the study was obtained from the village community leaders and the Institutional Ethics Committee of KEM Hospital Research Center, Pune.

### Statistical analysis

The purpose of our analysis was to determine the exposure variables over the life course significantly associated with cognitive outcomes at 12 years in the offspring. See tables 1 & 2 for a description of the dependent (outcome) and independent (exposure) variables used in this analysis. For maternal pregnancy exposures we used an average of measurements made at 18 and 28 weeks’ gestation. All variables were transformed to Z scores (by subtracting sample mean and dividing by the sample standard deviation for each variable). Height and weight for age Z scores at birth were calculated using the INTERGROWTH criteria.^35^ Height and BMI for age Z scores at 12 years were calculated according to the WHO reference 2007 using the WHO AnthroPlus software.^36^ We then performed multivariate linear regression for each cognitive score as dependent variable. The significant associations are interpreted as the SD change (standardized beta coefficient) in cognitive score for one SD change in the exposure variable.

**Table 1:** Maternal and paternal characteristics during pregnancy (1994-1996) and child characteristics at 12 years (2006-2008)

| Variables | N | Mean | Std. Deviation |
| --- | --- | --- | --- |
| **Maternal** |  |  |  |
| Age in years | 686 | 21.36 | 3.51 |
| Education in years | 664 | 5.73 | 3.93 |
| Pre pregnancy BMI (kg/m<sup>2</sup>) | 680 | 18.03 | 1.88 |
| Activity score | 684 | 69.92 | 23.11 |
| Total calorie intake | 684 | 1723.21 | 410.72 |
| Hb (gm%) | 678 | 11.11 | 1.72 |
| Fasting glucose (mg/dl) | 678 | 71.33 | 10.39 |
| Fasting insulin (pmol/L) | 675 | 46.08 | 339.61 |
| Cholesterol (mg/dl) | 678 | 169.96 | 33.32 |
| Triglycerides (mg/dl) | 678 | 113.2 | 33.5 |
| Vitamin B12 (pmol/L) | 674 | 145.24 | 66.99 |
| Folate (nmol/L) | 674 | 19.54 | 18.34 |
| Homocysteine (mmol/L) | 674 | 8.91 | 2.82 |
| Vitamin C (micromol/L) | 678 | 16.84 | 20.0 |
| Ferritin (mcg/L) | 678 | 15.8 | 14.59 |
| **Paternal** |  |  |  |
| Socio-economic status | 685 | 26.92 | 6.55 |
| education | 664 | 7.84 | 3.93 |
| BMI | 652 | 19.44 | 2.54 |
| **Birth** |  |  |  |
| Birth Wt for age Z score<sup>*</sup> | 610 | -1.56 | 0.93 |
| Birth Ht for age Z score<sup>*</sup> | 627 | -1.08 | 1.17 |
| Head circumference (cms) | 674 | 32.91 | 1.43 |
| **Child at 12 years** |  |  |  |
| Education in years | 681 | 5.88 | 1.28 |
| Ht for Age Z score<sup>#</sup> | 686 | -1.19 | 0.92 |
| BMI for Age Z score<sup>#</sup> | 686 | -1.71 | 1.18 |
| Head Circumference (cms) | 686 | 50.95 | 4.7 |
| Hb (gm%) | 686 | 12.78 | 1.32 |
| Glucose (mg/dl) | 683 | 87.02 | 7.14 |
| Fasting Insulin (pmol/L) | 687 | 5.79 | 3.29 |
| Cholesterol (mg/dl) | 686 | 130.95 | 23.84 |
| Triglycerides(mg/dl) | 686 | 58.63 | 22.12 |
| Vitamin B12 (pmol/L) | 681 | 207.89 | 87.58 |
| Folate (nmol/L) | 681 | 24.31 | 11.0 |
| Homocysteine (mmol/L) | 679 | 13.33 | 6.92 |
| Ferritin (mcg/L) | 652 | 22.09 | 15.02 |

\*Height and weight for age Z scores at birth calculated using the INTERGROWTH criteria
#Height and BMI for age Z scores at 12 years calculated according to WHO 2007 reference criteria.

**Table 2:** Neurocognitive performance scores in the children at 12 years

| Cognitive test | Mean<br>[Boys- 357] | S.D. | Mean<br>[Girls-329] | S.D. | P<br>value |
| --- | --- | --- | --- | --- | --- |
| **Color Progressive Matrices** | 28.5 | 7.4 | 27.5 | 7.2 | 0.07 |
| **Picture Completion** | 10.3 | 3.3 | 9.5 | 3.4 | **0.002\*** |
| **Digit span forward** | 4.9 | 1.3 | 4.9 | 1.3 | 0.9 |
| **Digit span backward** | 3.3 | 1.1 | 3.3 | 1.0 | 0.9 |
| **Color Trail making test A (time taken in secs)** | 79.9 | 37.1 | 74.1 | 26.1 | **0.02\*** |
| **Color Trail making test B (time taken in secs)** | 174.2 | 65.0 | 177.1 | 61.3 | 0.6 |
| **Auditory verbal learning test**<br>**(total learning score)** | 47.6 | 10.1 | 49.3 | 10.0 | **0.03\*** |
| **Block design (total correct score)** | 16.4 | 10.2 | 15.1 | 10.2 | 0.08 |

\*Significance at $P < 0.05$

In addition to using the linear regression model, we used BMA methods to estimate the coefficients of interest, and to account for model uncertainty. We move away from using any “benchmark” specification, as is often used in this literature because of the inherently open-ended nature of explanations for outcomes of interest such as child neurocognitive development, where the validity of one particular theory of neurocognitive development (e.g., in utero malnutrition) does not logically exclude other theories from also being relevant (e.g., socioeconomic background). In fact, the possibility of concurrent multiple exposures is implied by the life course model.

BMA, begins by defining a model space that is generated from the set of covariates for the dependent variable. A model is simply a particular permutation of the set of covariates. To address model uncertainty, heuristically, BMA assigns an evidentiary weight (i.e., the posterior model probability) to each model in the model space given the data, and then calculates the posterior distribution of the parameter of interest (e.g., the effect of in-utero maternal B12 concentrations on neurocognitive outcomes) by averaging across the set of models in the model space using these evidentiary weights (see Appendix B for technical details of the model). BMA analysis was performed using all the life course variables to provide a posterior mean and posterior inclusion probability (PIP) for every individual cognitive score.

As is standard in the literature, we report the posterior mean for each coefficient. The posterior mean is taken in the literature to be the model-averaged coefficient estimate for the effect. We also report the square root of the posterior variance as the corresponding standard error. Finally, we also report the posterior inclusion probability (PIP), the sum of the posterior probabilities of models that include that variable, for each covariate, which is a standard way to conduct inference for each regressor (supplementary material Appendix B). The standard way to conduct inference in the context of BMA is to reference the PIP for each regressor. Following the guidance provided by literature ^37,38^, we interpret a PIP < 50% as indicating a lack of evidence for an effect from that variable, a 50% < PIP < 75% as indicating weak evidence, a 75% < PIP < 95% as indicating positive evidence, a 95% < PIP < 99% as indicating strong evidence, and a 99% < PIP < 100% as indicating decisive evidence for an effect. Following standard practice^39^, we also report BMA posterior t statistics for coefficient estimates and interpret them in the classical sense. For the BMA results we report posterior means and posterior SD for associations with a PIP > 75%.

## Results

The mothers were on average 21 years old with a BMI of 18.03 ± 1.88 kg/m^2^ at the start of their pregnancy (Table 1). The average years of education was 5.73 ± 3.93 years and 35% mothers had not completed primary school education. The proportion of mothers who were anaemic (Hb<10 gm%) was 17.3%, average B12 level in mothers was 145.24 ± 66.99 pM, and 65% of mothers were B12 deficient (<150pM). The mean Ferritin level in the mothers during pregnancy was 15.8 ± 14.59 mcg/l. All mothers were folate replete and 29% mothers had hyper-homocysteinemia (>10 mmol/L). Fathers had an average BMI of 19.44 ± 2.54 kg/m^2^ with 4% being overweight, 20% had not completed primary school education. The new-born children weighed an average of 2.63 ± 0.37 kgs, 37 % of offspring were LBW and only 2 offspring were VLBW. 53% offspring were SGA and 10% were born preterm. At 12 years 18% of the children showed stunting and 94% were under-weight. The mean years of schooling was 5.88 ± 1.28 years. Only 5 out of the 686 children had dropped out of school. The cognitive test scores are provided in table-2. Boys performed better than girls on picture completion (visual attention) task (p=0.002) while girls performed better on TM B (attention) and AVLT (verbal memory) tasks (p=0.02 and 0.03 respectively). There was no difference in cognitive test scores between the offspring who were SGA compared to those born Average for Gestational Age (AGA) or between those born preterm or at term (supplementary table-1).

For conventional analysis, we present standardized beta coefficients, confidence intervals and significance level for the linear regression results for all the 8 neurocognitive development outcomes in Table 3 (A&B). Each column represents one outcome. The corresponding BMA results are shown in Table 4. All the results are summarised in Figure 2 (A&B). We show results in relation to major life-course determinants.

**Table 3A.**
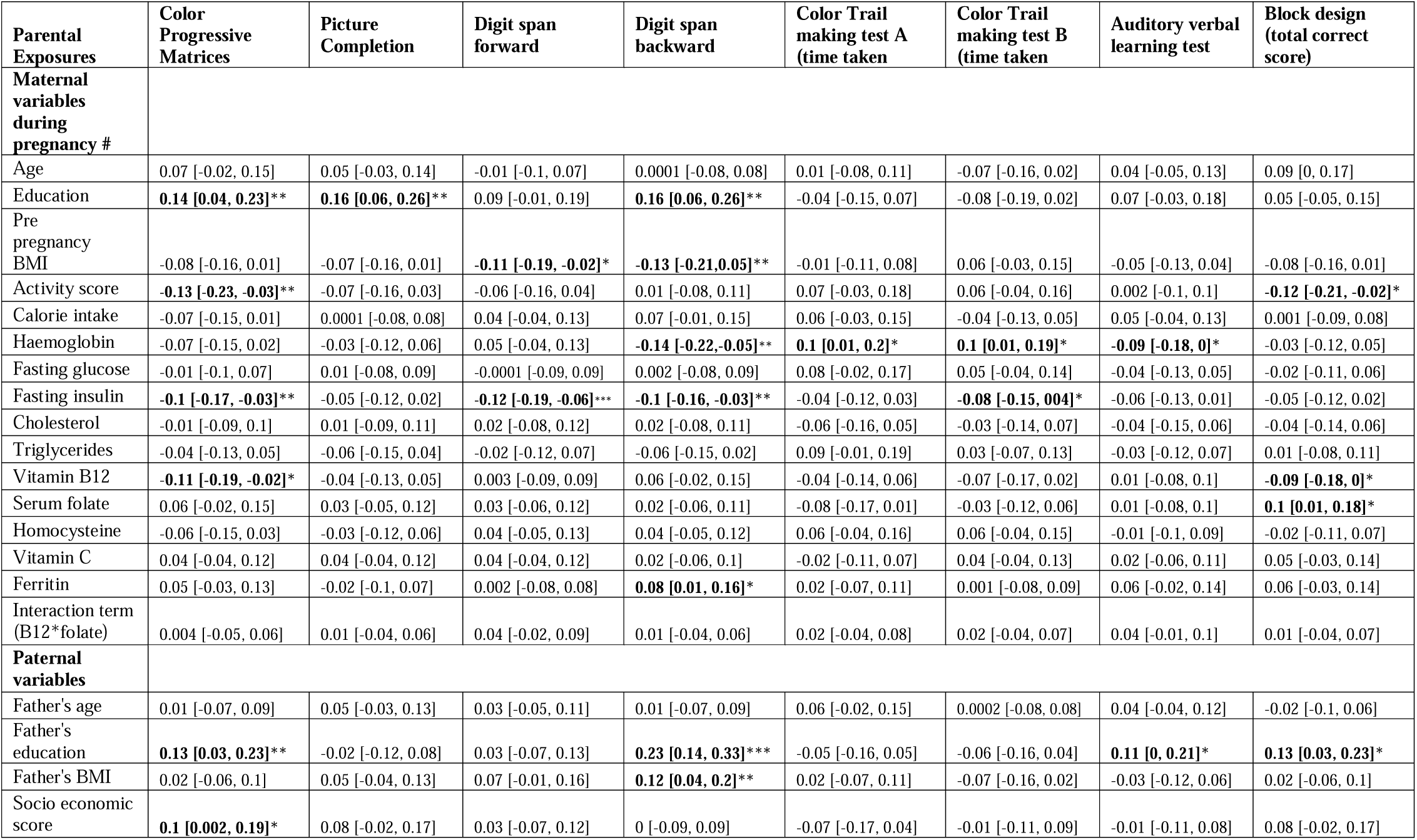
Results of Linear Regression analysis between parental exposures and offspring neurocognitive scores at age 12

**Table 3B.** Results of Linear Regression analysis between childhood variables and offspring neurocognitive scores at age 12

| Parental Exposures | Color Progressive Matrices | Picture Completion | Digit span forward | Digit span backward | Color Trail making test A (time taken) | Color Trail making test B (time taken) | Auditory verbal learning test | Block design (total correct score) |
| --- | --- | --- | --- | --- | --- | --- | --- | --- |
| **Child variables at birth** |  |  |  |  |  |  |  |  |
| Gender | 0.01 [-0.16, 0.18] | -0.1 [-0.27, 0.08] | 0.07 [-0.11, 0.24] | 0.11 [-0.06, 0.28] | **-0.22 [-0.41, -0.03]\*** | -0.01 [-0.19, 0.17] | **0.3 [0.12, 0.48]\*\*** | -0.02 [-0.2, 0.16] |
| Head circumference | -0.02 [-0.14, 0.1] | **0.14 [0.02, 0.27]\*** | 0.11 [-0.02, 0.24] | 0.08 [-0.05, 0.2] | -0.11 [-0.24, 0.03] | -0.05 [-0.18, 0.08] | 0.04 [-0.09, 0.17] | 0.09 [-0.04, 0.22] |
| Length | 0.1 [-0.03, 0.22] | 0.07 [-0.06, 0.19] | -0.07 [-0.2, 0.05] | -0.04 [-0.16, 0.08] | -0.08 [-0.21, 0.06] | -0.06 [-0.19, 0.07] | -0.02 [-0.15, 0.11] | 0.02 [-0.11, 0.15] |
| Birth weight | -0.002 [-0.14, 0.14] | -0.14 [-0.28, 0.01] | 0.03 [-0.12, 0.18] | 0.02 [-0.12, 0.16] | 0.11 [-0.05, 0.27] | **0.2 [0.05, 0.36]\*\*** | 0.001 [-0.15, 0.15] | -0.07 [-0.21, 0.08] |
| **Child variables at 12 years** |  |  |  |  |  |  |  |  |
| Height | **0.19 [0.04, 0.34]\*** | 0.03 [-0.12, 0.18] | **0.19 [0.04, 0.34]\*** | **0.15 [0.01, 0.29]\*** | -0.16 [-0.32, 0.01] | -0.14 [-0.3, 0.01] | -0.01 [-0.16, 0.15] | **0.23 [0.08, 0.38]\*\*** |
| Weight | -0.1 [-0.26, 0.06] | 0.14 [-0.02, 0.31] | -0.09 [-0.26, 0.08] | **-0.18 [-0.34, -0.02]\*** | 0.16 [-0.02, 0.34] | 0.01 [-0.16, 0.19] | -0.02 [-0.2, 0.15] | -0.003 [-0.17, 0.17] |
| Head circumference | 0.07 [-0.01, 0.14] | 0.02 [-0.05, 0.1] | -0.01 [-0.09, 0.06] | 0.004 [-0.07, 0.08] | -0.001 [-0.08, 0.08] | 0.03 [-0.04, 0.11] | -0.01 [-0.08, 0.07] | -0.04 [-0.12, 0.03] |
| Haemoglobin | -0.06 [-0.16, 0.05] | -0.07 [-0.18, 0.04] | 0.05 [-0.06, 0.16] | 0.003 [-0.1, 0.11] | 0.02 [-0.1, 0.14] | 0.05 [-0.07, 0.16] | -0.02 [-0.13, 0.1] | -0.04 [-0.15, 0.07] |
| Fasting glucose | -0.02 [-0.1, 0.07] | -0.05 [-0.13, 0.04] | -0.01 [-0.09, 0.08] | -0.06 [-0.14, 0.02] | -0.02 [-0.11, 0.07] | 0.003 [-0.08, 0.09] | -0.01 [-0.09, 0.08] | -0.02 [-0.11, 0.06] |
| Fasting insulin | 0.04 [-0.06, 0.14] | -0.03 [-0.13, 0.07] | -0.05 [-0.14, 0.05] | 0.06 [-0.03, 0.16] | -0.02 [-0.13, 0.09] | -0.003 [-0.11, 0.1] | -0.01 [-0.11, 0.09] | **-0.12 [-0.22, -0.02]\*** |
| Cholesterol | -0.01 [-0.11, 0.08] | 0.02 [-0.08, 0.11] | 0.001 [-0.1, 0.1] | -0.06 [-0.15, 0.03] | 0.05 [-0.06, 0.16] | 0.02 [-0.08, 0.12] | 0.02 [-0.08, 0.12] | 0.01 [-0.09, 0.1] |
| Triglycerides | 0.01 [-0.09, 0.11] | -0.07 [-0.17, 0.02] | 0.05 [-0.05, 0.15] | -0.01 [-0.1, 0.09] | **-0.12 [-0.23, -0.01]\*** | 0.01 [-0.1, 0.11] | -0.01 [-0.11, 0.09] | -0.02 [-0.12, 0.08] |
| Vitamin B12 | -0.05 [-0.14, 0.03] | 0.02 [-0.07, 0.1] | -0.07 [-0.16, 0.02] | 0.01 [-0.07, 0.1] | -0.03 [-0.13, 0.06] | 0.001 [-0.09, 0.09] | **0.09 [0, 0.18]\*** | -0.01 [-0.1, 0.08] |
| Folate | **-0.08 [-0.16, 0]\*** | -0.03 [-0.11, 0.05] | -0.07 [-0.15, 0.01] | -0.06 [-0.13, 0.02] | 0.07 [-0.02, 0.15] | 0.06 [-0.03, 0.14] | -0.05 [-0.14, 0.03] | -0.07 [-0.15, 0.01] |
| Homocysteine | -0.06 [-0.15, 0.04] | 0.08 [-0.01, 0.18] | -0.04 [-0.13, 0.06] | -0.07 [-0.17, 0.02] | 0.05 [-0.05, 0.16] | 0.02 [-0.08, 0.12] | -0.003 [-0.1, 0.1] | 0.02 [-0.08, 0.12] |
| Ferritin | 0.02 [-0.06, 0.11] | -0.01 [-0.09, 0.07] | 0.02 [-0.07, 0.1] | 0.04 [-0.04, 0.12] | -0.02 [-0.11, 0.07] | -0.06 [-0.15, 0.03] | 0.04 [-0.05, 0.13] | -0.01 [-0.09, 0.08] |

Significance at \*\*\*p<0.001, \*\*p<0.01, \*p<0.05.
Each column represents a linear regression with the column heading as its dependent variable. The $\beta$ -coefficients are included for each variable for each regression, with the 95% confidence interval in parentheses. All the regressions included maternal, paternal, child at birth as well as child at 12-years variables. # - The maternal variables are the mean of measurements made at 18 and 28-week pregnancy.

**Table 4.** Results of Bayesian analysis

| Parental Exposures | Color Progressive Matrices | Picture Completion | Digit span forward | Digit span backward | Color Trail making test A (time taken) | Color Trail making test B (time taken) | Auditory verbal learning test | Block design (total correct score) |
| --- | --- | --- | --- | --- | --- | --- | --- | --- |
| **Maternal variables during pregnancy #** |  |  |  |  |  |  |  |  |
| Age | --- | --- | --- | --- | --- | --- | --- | --- |
| Education | 0.14(0.08)* | 0.19(0.04)*** | 0.14(0.06)* | 0.15(0.06)* | --- | --- | --- | --- |
| Pre pregnancy BMI | --- | --- | --- | -0.12(0.06)* | --- | --- | --- | --- |
| Activity score | --- | --- | --- | --- | --- | --- | --- | --- |
| Calorie intake | --- | --- | --- | --- | --- | --- | --- | --- |
| Haemoglobin | --- | --- | --- | -0.1(0.06)* | --- | --- | --- | --- |
| Fasting glucose | --- | --- | --- | --- | --- | --- | --- | --- |
| Fasting insulin | --- | --- | -<br>0.13(0.04)** | --- | --- | --- | --- | --- |
| Cholesterol | --- | --- | --- | --- | --- | --- | --- | --- |
| Triglycerides | --- | --- | --- | --- | --- | --- | --- | --- |
| Vitamin B12 | --- | --- | --- | --- | --- | --- | --- | --- |
| Serum folate | --- | --- | --- | --- | --- | --- | --- | --- |
| Homocysteine | --- | --- | --- | --- | --- | --- | --- | --- |
| Vitamin C | --- | --- | --- | --- | --- | --- | --- | --- |
| Ferritin | --- | --- | --- | --- | --- | --- | --- | --- |
| Interaction term (B12*folate) | --- | --- | --- | --- | --- | --- | --- | --- |
| **Paternal variables** |  |  |  |  |  |  |  |  |
| Father's age | --- | --- | --- | --- | --- | --- | --- | --- |
| Father's education | 0.16(0.07)* | --- | --- | 0.24(0.05)*** | --- | --- | --- | 0.19(0.05)** |
| Father's BMI | --- | --- | --- | --- | --- | --- | --- | --- |
| Socio economic score | --- | --- | --- | --- | --- | --- | --- | --- |

|  | Color<br>Progressive<br>Matrices | Picture<br>Completion | Digit span<br>forward | Digit span<br>backward | Color Trail<br>making test<br>A (time<br>taken) | Color Trail<br>making test<br>B (time<br>taken) | Auditory<br>verbal<br>learning test | Block design<br>(total correct<br>score) |
| --- | --- | --- | --- | --- | --- | --- | --- | --- |
| **Child Exposures** |  |  |  |  |  |  |  |  |
| **Child variables at birth** |  |  |  |  |  |  |  |  |
| Gender | --- | --- | --- | --- | --- | --- | 0.27(0.11)* | --- |
| Head circumference | --- | --- | --- | --- | --- | --- | --- | --- |
| Length | --- | --- | --- | --- | --- | --- | --- | --- |
| Birth weight | --- | --- | --- | --- | --- | --- | --- | --- |
| **Child variables at 12 years** |  |  |  |  |  |  |  |  |
| Height | 0.14(0.06)* | --- | --- | --- | --- | -0.11(0.07)* | --- | 0.23(0.05)*** |
| Weight | --- | --- | --- | --- | --- | --- | --- | --- |
| Head circumference | --- | --- | --- | --- | --- | --- | --- | --- |
| Haemoglobin | --- | --- | --- | --- | --- | --- | --- | --- |
| Fasting glucose | --- | --- | --- | --- | --- | --- | --- | --- |
| Fasting insulin | --- | --- | --- | --- | --- | --- | --- | --- |
| Cholesterol | --- | --- | --- | --- | --- | --- | --- | --- |
| Triglycerides | --- | --- | --- | --- | --- | --- | --- | --- |
| Vitamin B12 | --- | --- | --- | --- | --- | --- | --- | --- |
| Folate | --- | --- | --- | --- | --- | --- | --- | --- |
| Homocysteine | --- | --- | --- | --- | --- | --- | --- | --- |
| Ferritin | --- | --- | --- | --- | --- | --- | --- | --- |

\*\*\* PIP>99%, \*\* >95%PIP<99%, \* >75% PIP <95%. . Each column represents a BMA regression with the column heading as its dependent variable. The posterior mean are shown in each cell of the table, and the square root of posterior variances are in parentheses. Only robust predictors (PIP>75%) are depicted in the table.

**Figure 2:**
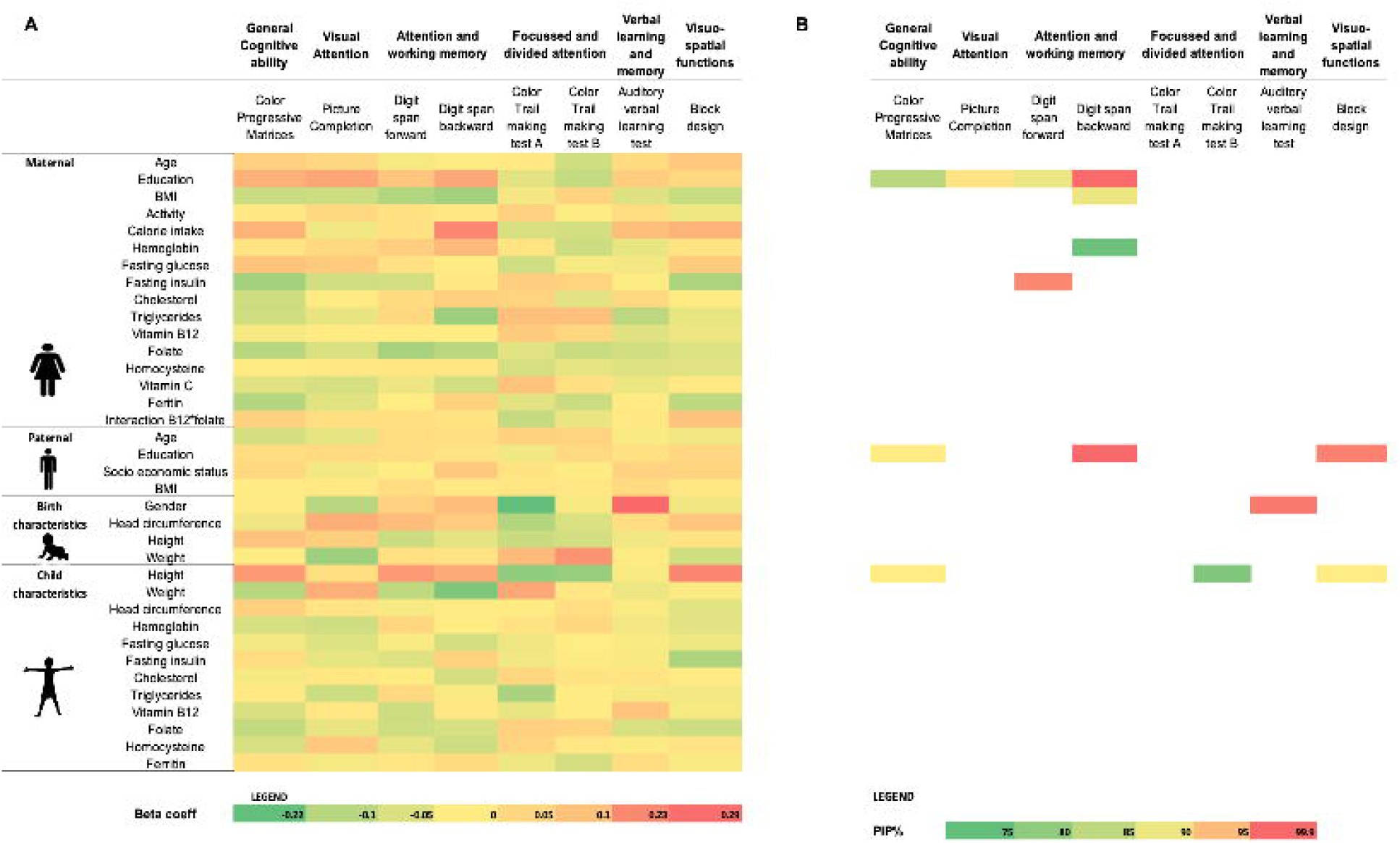
(A) Represents linear regression beta coefficients for associations between life-course variables and 12 year neurocognitive scores in the offspring. Figure (B) represents Bayesian Model Averaging (BMA) Posterior Inclusion Probability (PIP) % for robust predictors.

### Maternal Condition During Pregnancy

We also found some evidence that maternal condition during pregnancy was associated with child’s neurocognitive outcomes. Higher maternal BMI was associated with poorer attention (lower scores on DF and DB β = -0.11 [95% CI -0.19, -0.02], β = -0.13 [95% CI -0.21, -0.05]) respectively), higher physical activity scores was associated with poorer general cognitive ability (lower CPM scores β = -0.13 [95% CI -0.23, -0.03]) and higher fasting insulin levels with lower performance on tasks of memory and attention (DF β = -0.12 [95% CI -0.19, -0.06], and DB β = -0.1 [95% CI -0.16, -0.03]) and lower general cognitive ability (CPM β = -0.1 [95% CI -0.17, -0.03]). We observed small effects for higher maternal folate on better visuo-spatial ability (higher BD score β = 0.1 [95% CI 0.01, 0.18]) and higher maternal ferritin on better attention (higher DB score β = 0.08 [95% CI 0.01, 0.16]). Higher maternal haemoglobin was associated with poorer attention and verbal memory and higher maternal B12 with poorer general cognitive ability. The interaction term (maternal folate X B12) was not significantly associated with any of the cognitive outcomes.

Nevertheless, with two exceptions, our BMA results would suggest that the above findings are not robust. The exceptions were for higher mother’s fasting insulin levels being negatively associated with attention (DF scores 98.07% PIP) and higher pre-pregnancy BMI with lower DB scores (90% PIP).

### Child’s Birth Characteristics and Subsequent Development

Girls performed significantly better than boys on attention and memory (longer TM A time β = -0.22 [95% CI -0.41, -0.03] and AVLT test scoresβ= 0.30 [95% CI 0.12, 0.48]). Larger head circumference at birth was associated with significantly better visual attention (PC scores β = 0.14 [95% CI 0.02, 0.27]), and higher birth weight with poorer focussed attention (TM B time β = 0.2 [95%CI 0.05, 0.36]). Higher height-for-age at age 12 was associated with better global neurocognitive function (CPM scores β = 0.19 [95% CI 0.04, 0.34]), attention (DF scores β = 0.14 [95% CI 0.04, 0.34]), DB scores β = 0.15 [95% CI 0.01, 0.29]) and visuo-spatial abilities (BD scores β = 0.23 [95% CI 0.08, 0.38]). In addition, higher fasting insulin levels at 12 years were associated with poorer visuo-spatial abilities (lower BD scores β = -0.12 [95% CI -0.22, -0.02]) and higher triglyceride levels with better attention (TM A times β = -0.12 [95% CI -0.23, -0.01]). Micronutrient concentrations in the child were also significantly associated with neurocognitive performance: Higher B12 levels with better verbal learning (AVLT scores β = 0.09 [95% CI 0, 0.18]) and higher folate levels with worse general cognitive ability (CPM scores β = -0.08 [95% CI -0.16, 0]).

BMA results suggest that only gender differences as well as the higher height-for-age at age 12 remain robustly associated with our outcome variables BD scores (99.8% PIP), CPM scores (92.1% PIP) and TM B time (78.3% PIP).

### Parental education and family socioeconomic status

The most consistent finding across the set of child’s neurocognitive outcomes is the positive association with parental education and their SES, especially the former. Neurocognitive performance in all the tests, with the exception of measures of attention (TM A and TM B), showed strong positive associations.

We found that parental education and SES were independently associated with global neurocognitive ability (CPM, β=0.13 [95% CI 0.03, 0.23] and β=0.1 [95% CI 0.002, 0.19] respectively). Father’s education was more strongly associated with tests ofattention and working memory (DB, β=0.23 [95% CI 0.14, 0.33]) than mother’s education (β=0.16 [95% CI 0.06, 0.26]). Father’s education was associated with better performance on visuo-spatial abilities and memory (BD β = 0.13 [95% CI 0.03, 0.23] and AVLT β = 0.11 [95% CI 0, 0.21]) while mothers education correlated with higher visual attention (PC) (β = 0.16 [95% CI 0.06, 0.26]).

These findings were largely affirmed to be robust by our BMA exercises. Thus, father’s education was found to be a robust determinant of general cognitive ability(CPM 92.22% PIP), visuo-spatial abilities (BD 98.5% PIP), and attention (DB 99.99% PIP), while mother’s education for general cognitive ability (CPM 84.5% PIP), and attention (DB 92.69% PIP, DF 89.83% PIP, PC 99.86% PIP).

## Discussion

Our BMA analysis supports a robust role for parental education, maternal pre pregnancy BMI and pregnancy insulin concentrations, and child’s height in influencing neurocognitive outcomes in the offspring at 12 years of age. Associations of maternal nutritional status during pregnancy (folate and ferritin levels) and child’s birth measurements (head circumference) were less robustly associated. These results are in line with the recent literature that suggests that parental influence and family background play a vital role in children’s neurocognitive development ^40,41^.

Parental education stood out as the most robust predictor on offspring cognitive outcome. This may influence the neurocognitive outcomes through many pathways. The effects of parental education on offspring cognition may operate through a more stimulating home environment and better opportunities for education^41,42^. This is supported by a two generation behavioral intervention study that showed that parental training to promote opportunities for children at school and home resulted in better school achievement and reduced behavioral problems in their offspring^43^. The benefits of such improved home environment continued into the next offspring who also showed better early child developmental functioning. The Swedish sibling adoption study demonstrates the role of socio-economic environment on neurocognitive ability in the most definitive way^44^. This study examined the intelligence of sibling pairs at 18 years of age, where one sibling was reared by the biological parents and the other by adoptive parents who had a higher parental education. The results showed that the adoptive sibling scored 4.41 IQ points higher than the non-adoptive sibling. Each additional unit of education of the rearing parent, was associated with a 1.94 unit increase in IQ in the offspring.

Among the early developmental factors, starting from preconception we observed that higher pre-pregnancy BMI was associated with lower cognitive scores in the offspring. Findings from developed countries show an association between maternal obesity in pregnancy and a 2 point lower IQ in the offspring at age 7 years.^45^ A meta-analysis of 32 studies found that maternal pre-pregnancy overweight or obesity was associated with higher odds of adverse neurodevelopmental outcomes in the offspring^46^. However, mothers in our study had a low BMI (average pre-pregnancy BMI ∼ 18) which makes it difficult to interpret the inverse association. One possible explanation may be related to the relatively high adiposity in Indians,^47^ so that the inverse association may reflect an effect of a more healthy body composition of the mother.^48^ Similarly, association of higher maternal fasting insulin with poor cognitive outcomes in the offspring is not easy to explain because insulin concentrations were quite low and mothers had a normal glucose tolerance. Overall, we interpret our results to indicate that maternal size, body composition and metabolism in pregnancy are important determinants of offspring brain development and its cognitive performance but this needs further exploration in future studies.

Micronutrients vitamins B12, folate, C and D, pyridoxine and ferritin are cofactors in important celluar processes (DNA synthesis, epigenetic regulation, cell cycle regulation and energy metabolism) that impact neurodevelopment. Systematic reviews report associations between lower maternal concentrations of these micronutrients during pregnancy and enhanced risk for various adverse neurodevelopmental outcomes such as neural tube defects, autism and poorer cognitive performance in the offspring^49,50^. We observed only small effects for maternal folate and ferritin on visuo-spatial ability and attention respectively at 12 years, which were not robust under BMA. We had earlier demonstrated in a small pilot study that offspring exposed to lowest decile of maternal B12 concentrations in-utero had poorer cognitive performance at 9 years of age compared to offspring in the highest decile of maternal B12 concentrations during pregnancy.^15^ We did not observe such associations in the current analysis. This may suggest that the previous results were “fragile” (in the sense of Leamer (1983)^51^) and not robust to model uncertainty. However, recently we reported findings from a randomized controlled trial performed in the same cohort.^52^ Pre-conceptional B12 supplementation improved cognitive and language outcomes in the offspring at 2-4 years of age supporting a causal role for maternal vitamin B12 status in offspring neurodevelopment. It would be interesting to see if this effect lasts at a later age.

Among birth characteristics, being born with a low birth weight is consistently associated with adverse cognitive outcomes. A meta-analysis of 35 studies on neurocognitive outcomes in low birth weight children showed moderate to severe deficits in attention, executive functions and academic achievement^53^. In another meta-analysis of 13 studies, being born with VLBW or Very Pre-Term (VPT – gestation <32 weeks) was associated with a 12 point lower IQ score in adulthood^54^. A prospective hospital based study, in infants born with VLBW from Pune India, found preterm children who were small for gestational age to have lower IQ scores in adulthood ^55^. In the current study 37% of offspring were LBW, two (0.3%) were VLBW while 10% were born preterm. There were no differences in cognitive outcomes in offspring born SGA or AGA and between those born preterm or term There were modest associations between higher birth weight and lower head circumference with poorer scores on attention tasks. However none of the birth characteristics were robust predictors under BMA.

Among the post-natal developmental influences, we found stunting to be the most consistent predictor of poorer cognitive performance. Child’s height at 12 years was associated with many cognitive outcomes including generalised cognitive ability, attention and visuo-spatial ability. A metanalysis of 68 studies found a 0.22 SD increase in cognitive function score per SD increase in height for age Z scores at age 5-11 years ^56^. Other child anthropometric measures were not robustly associated under BMA.

Our BMA results generally find that only a few maternal biomarkers and initial conditions are significant, and that the most important factors determining children’s neurocognitive outcomes relate to parental education and socio-economic background. The pathways for this influence may be multiple. Parental education will influence their nutrition, metabolism and behaviour in many ways from before conception, during pregnancy and continue to influence the home environment (including diet, habits, intellectual stimulation and opportunities etc) in subsequent years. The much investigated nutritional and other biological exposures appear to be embedded in parental education. Developmental biology has stressed the most prominent window for such an influence to be early in life (pre and periconceptional period, pregnancy and first two years of life) which is popularised by the term “first 1000 days”. An additional consideration is the ability of educated parents to take remedial action in response to a faltering signal. All these will influence the performance of the developing nervous system^57–59^.

The strength of our study is the novelty of using a BMA on life course exposure variables to identify robust predictors of cognitive outcomes in childhood. The data represented in this study traces exposures over the life course from pre-pregnancy and pregnancy to12 years of age of the offspring. These observations are unique and allow us to test the DOHaD paradigm. These longitudinal exposures encompass socio-economic transition and impact of various Government of India schemes to improve childhood nutrition and development over the life course of these children (from 1993 to 2008). We acknowledge that the exposures will change with time. We continue to follow this cohort and performed neurocognitive assessments and brain imaging at 24 years of age which will allow us to examine more recent exposures and their impact on longer term cognitive outcomes into adulthood^60^. An additional strength of our study is that we used a comprehensive, culturally relevant measure to assess socioeconomic status of rural Indian parents. Family income alone is considered as an unreliable estimate of socioeconomic status ^61^ and educational attainment and occupational status are known to be better indicators ^62^. One limitation of our study is that, because of unavailability of data, we were not able to specifically assess family home environment and parent child interactions which are known to moderate associations of SES with child neurocognitive performance. Finally, being an observational, non-randomized study we cannot be certain about the causality of associations.

## Conclusion

In this paper, we investigated the linkages that life course exposures to various biological and socio-economic factors have on neurocognitive performance at age 12 using data from the Pune Maternal Nutrition Study (PMNS). The exposures included socioeconomic background as well as a range of biometric measures for the mother and the child, from birth until the time at which the neurocognitive tests were taken. In our analysis, we explicitly account for the important issue of model uncertainty in regression exercises by exploiting the BMA methodology.

Across the measures of neurocognitive abilities, once we account for model uncertainty, we generally find overwhelming importance for the role of parental education in determining a child’s outcomes at age 12. The associations with child’s height indicate that remedial interventions in the post-natal life may have important and significant associations with neurocognitive development. Though nutritional deficiencies in-utero or size at birth have long-term influence, those effects are potentially reversed by post-birth remediation. Coupled with the success of our vitamin B12 intervention trial, current results inform public health policy in India to consider social interventions in children born into families with low socio-economic status to improve human capital, over and above the already operational nutritional programs.

## Supporting information

There was no difference in cognitive test scores between the offspring who were SGA compared to those born Average for Gestational Age (AGA) or betwee

## Data Availability

All data produced in the present study are available upon reasonable request to the authors

## Acknowledgement

The authors thank Mrs. Aboli Bhalerao for her help with data management and statistical analysis.

