## Supplementary material for "Robust Determinants of Neurocognitive Development in Children: Evidence from the Pune Maternal Nutrition Study (PMNS)": There was no difference in cognitive test scores between the offspring who were SGA compared to those born Average for Gestational Age (AGA) or betwee

**Supplementary table 1: Neurocognitive performance of offspring in SGA vs AGA and preterm vs term**

| **Cognitive scores at 12 years** | SGA (n=344)  Median  (Q1, Q4) | AGA (n=297)  Median  (Q1, Q4) | P - Value | Preterm (n=67)  Median  (Q1, Q4) | Term (n=601)  Median  (Q1, Q4) | P - value |
| --- | --- | --- | --- | --- | --- | --- |
| Color Progressive Matrices | 30  (24, 33.8) | 31  (25, 34) | 0.14 | 29  (23, 33) | 30  (25, 34) | 0.11 |
| Picture Completion | 10  (7, 12) | 10  (8, 12) | 0.28 | 10  (6, 12) | 10  (8, 12 | 0.14 |
| Digit span forward | 5  (4, 6) | 5  (4, 6) | 0.40 | 4  (4, 5) | 5  (4, 6) | 0.18 |
| Digit span backward | 3  (3, 4) | 3  (3, 4) | 0.15 | 3  (2, 4) | 3  (3, 4) | 0.22 |
| Color Trail making test A (time taken in secs) | 71  (57, 90) | 72  (59, 90) | 0.28 | 78  (57, 96) | 70  (58, 89) | 0.29 |
| Color Trail making test B (time taken in secs) | 168.5  (135, 208.5) | 171  (137, 205) | 0.64 | 167  (129,206) | 170  (135, 207) | 0.65 |
| Auditory verbal learning test  (total learning score) | 49  (42, 56) | 49  (43, 55.5) | 0.58 | 49  (42, 55) | 49  (43, 56) | 0.52 |
| Block design (total correct score) | 14  (6, 22) | 14  (6, 24.5) | 0.07 | 10  (6, 21) | 14  (6, 23.5) | 0.11 |

**Appendix A: Neurocognitive assessment at 12 years in the PMNS**

| **Cognitive test** | **Domain tested** | **Administration** | **Score** | **Interpretation** |
| --- | --- | --- | --- | --- |
| Colored Progressive Matrices | It is a measure of global cognitive function designed for young children which measures ability to think and perceive. | The child was presented with a series of patterns with a missing piece and had to identify the pattern from six options provided. The test consists of 36 items in 3 sets, each set containing 12 items. Test-retest reliability of CPM shows encouraging evidence of its stability in various cultures. | A score was allotted for every pattern recognized and a total score was generated. Time taken was also recorded.  Raw scores and Percentile Rank (PR) / Total Quotient (TQ) were calculated as per standard norms available. | Higher score indicates better global cognitive function. |
| Picture completion | Visuo-Conceptual ability. Picture completion is also a subtest from Weschler’s intelligence scale for children. It measures visuo-conceptual ability, which is the ability to combine disconnected, vague visual stimuli into a meaningful whole by matching them with their long-term representations | The child was shown a set of pictures with a part missing. They had to identify the missing part within a given time limit | Total number of correct responses provided within the time limit was recorded. Raw scores and PR/TQ were calculated as per standard norms available. | Higher score indicates better visual conceptual ability |
| Digit Span -Forward and Backward | Working memory. This test is a measure for working memory and is taken from Weschler’s intelligence scale for children. | It consists of two parts, forward and backward. Strings of numbers in increasing length are given which the subject has to recall in forward direction for digit forward and in backward direction for digit backward. The longest digit string recalled is considered as the span. The child had to repeat a series of numbers called out by the tester and had to recall them in the same order (Forward).The span of numbers increased after correct responses were given on the span . For digit span backward, the child had to recall numbers in a reverse order. | Forward and backward scores were calculated individually as per correct responses. Raw scores and PR/TQ were calculated as per standard norms available. | Higher score indicates better working memory |
| Color Trail Making A & B | It is a measure of Focused attention, also measures conceptual tracking of mental ability. | **Colour Trail making A:** The test contained pink and yellow circles containing numbers from 1-25. In the first part, the child had to connect the circles containing the numbers, in ascending order. When connecting the numbers, the child was instructed not to lift the pencil/pen and not to draw over already constructed lines.  **Colour Trail making B:** In the second part the child again had to connect rapidly the numbered circles in sequence, but alternates between pink and yellow. | The time taken to complete the test and number of errors were recorded. Raw scores and PR/TQ were calculated as per standard norms available. | Lower time and errors taken indicates better Focused attention |
| Auditory Verbal Learning Test (AVLT) | Verbal learning and memory | The child was read out a list of 15 words and was asked to recall the words. The list (list A) was initially read out 5 times and recall was tested after each reading. After this, a second list was read out (list B) and recall was tested. Following the recall of list B, recall of list A was immediately tested again and after a gap of 20 minutes. | The child was scored based on number of correct answers for both immediate recall and delayed recall. Raw scores and PR/TQ were calculated as per standard norms available. | Higher score indicates better verbal learning and memory |
| Block design | Visuo-spatial ability. This is a subtest from Weschsler’s intelligence scale for children. It measures visuo-spatial skills i.e., the ability to break down spatial forms into their constituent elements. | The subject is given red and white blocks. The task is to use the blocks to construct the replicas of designs given on cards. Designs are given in order of increasing difficulty.  Child had to view a design and recreate that design using 3-D blocks by assembling blocks. Multiple designs of complexity were administered in sequence. | A score was allotted for every design completed in a specified time limit for that design. A total score was then computed. Raw scores and PR/TQ were calculated as per standard norms available. | Higher score indicates better visuo-spatial ability |

This battery of neuropsychological tests was developed to measure the neuro cognitive function in children. All the tests were age specific and Indian normative percentiles and cut-off scores were used for scoring. For WISC (Malin’s- Indian version) was used. TQ/PR were generated from Indian standard norms available from NIMHANS and Malins- Indian Version as applicable for each test.

**Appendix B: Statistical Analysis**

We refer the reader to Raftery et al. (1997)^1^ for a detailed discussion of the econometrics behind BMA. BMA allows researchers to address the issue of potential “fragility” of estimators under model uncertainty that potentially results from “theory open-endedness”. The notion of “theory open-endedness” was introduced by Brock and Durlauf (2001) ^2^ in the context of economic growth who argued that this problem potentially renders the estimates of coefficients of interest to be “fragile” in the sense of Leamer (1978) ^3^. By fragility, they mean that the estimated effect could change dramatically in magnitude, lose its statistical significance, or, even switch signs depending on which other control variables are included or excluded in the regression equation. Such fragility renders policy statements based upon these findings uncompelling.

Formally, let the effect of interest be $\beta_{z}$. The posterior distribution of this parameter is

$P\left( \beta_{z} | D \right)=\sum_{k=1}^{K} P\left( \beta_{z} | M_{k},D \right)P(M_{k}|D)$	(3.1)

where
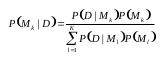
(3.2)

and where
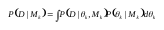
(3.3)

where $\theta_{k}$ is the vector of parameters of $M_{k}$, $P(\theta_{k}|M_{k})$ is the prior density of $\theta_{k}$ under the model $M_{k}$, $P(D|\theta_{k},M_{k})$ is the marginal likelihood, and $P(M_{k})$ is the prior probability that $M_{k}$is the true model.

With this information, the posterior mean and variance can be determined as follows:

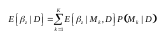
(3.4)
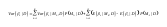
 (3.5)

In practice, the model space may be very large since it consists of 2^k^ models where k is the number of covariates such that averaging across the entire set of models is computationally infeasible. Researchers therefore typically employ a stochastic search algorithm that takes the process to regions of high posterior probabilities in the model space and to average across those models. BMA also requires that we specify a prior distribution across models that capture prior knowledge about the plausibility of models. It is standard in this literature to employ non-informative model priors to capture agnosticism across models in the model space. Specifically, we set the model prior to be uniform, where the prior probability of a covariate being in the true model is 0.5 (agnostic).

BMA also requires us to specify priors over parameters. Following standard choices from literature, we report results for g priors that are estimated using Empirical Bayes^4,5^. In terms of the settings for the MCMC stochastic search algorithm, we use a burn-in phase of 100,000 draws, and then calculate posterior probabilities based on 1 million successive draws.  After 1 million draws, the correlation of posterior model probabilities was over 0.99 across the exercises indicating that the 20,000 most successful models have converged over the million draws.
